# Studies of parenchymal texture added to mammographic breast density and risk of breast cancer: a systematic review of the methods used in the literature

**DOI:** 10.1101/2021.11.16.21266374

**Authors:** Akila Anandarajah, Yongzhen Chen, Graham A. Colditz, Angela Hardi, Carolyn Stoll, Shu Jiang

## Abstract

This systematic review aimed to assess the methods used to classify mammographic breast parenchymal features in relation to prediction of future breast cancer including the time from mammogram to diagnosis of breast cancer, and methods for the identification of texture features and selection of features for inclusion in analysis. The databases including Medline (Ovid) 1946-, Embase.com 1947-, CINAHL Plus 1937-, Scopus 1823-, Cochrane Library (including CENTRAL), and Clinicaltrials.gov. were searched through October 2021 to extract published articles in English describing the relationship of parenchymal texture features with risk of breast cancer. Twenty-eight articles published since 2016 were included in the final review. Of these, 7 assessed texture features from film mammograms images, 3 did not report details of the image used, and the others used full field mammograms from Hologic, GE and other manufacturers. The identification of parenchymal texture features varied from using a predefined list to machine-driven identification. Reduction in number of features chosen for analysis in relation to cancer incidence then varied across statistical approaches and machine learning methods. The variation in approach and number of features identified for inclusion in analysis precluded generating a quantitative summary or meta-analysis of the value of these to improve predicting risk of future breast cancers. This updated overview of the state of the art revealed research gaps; based on these, we provide recommendations for future studies using parenchymal features for mammogram images to make use of accumulating image data, and external validation of prediction models that extend to 5 and 10 years to guide clinical risk management. By following these recommendations, we expect to improve risk classification and risk prediction for women to tailor screening and prevention strategies to level of risk.

## Introduction

Evolving technology from film mammograms to digital images has changed the sources of data and ease of access to study a range of summary measures from breast mammograms and risk of breast cancer.^1,2^ These include more extreme measures of density and also measures of breast texture features. In particular, as women have repeated mammograms as part of a regular screening program,^3-5^ access to repeated images including changing texture features has become more feasible in real time for risk classification and counseling women for their risk management.^6-8^ Using these features may facilitate improvement in risk classification^9^ and hence more fully support precision prevention for breast cancer.^8,10^

The leading measure for risk categorization extracted from mammograms is breast density.^11,12^ This is now widely used and reported with many states requiring return of mammographic breast density measures to women as part of routine screening. Mammographic breast density is a strong reproducible risk factor for breast cancer across different approaches used to measure it (clinical judgement or semi/automated estimation), and across regions of the world.^11^ Other texture features within mammograms have been much less frequently studied for risk stratification and risk prediction. However, growing access to the large data from digital mammograms encourages exploration of additional texture features, complimentary to mammogram density, to assess risk of subsequent breast cancer.^13,14^

While patterns of breast parenchymal complexity, formed by the x-ray attenuation of fatty, fibroglandular, and stromal tissues, are known to be associated with breast cancer,^15,16^ mammogram density only aims to measure the relative amount of fibroglandular tissue in the breast,^17^ which limits the ability to fully capture heterogeneity between patients in the breast tissue. In 2016 Gastounioti and colleagues summarized the literature at that time to classify approaches to parenchymal texture classification: 1) grey-level features – skewness; kurtosis; entropy; and sum intensity, 2) co-occurrence features – entropy; inertia; difference moment; and coarseness, 3) run-length measures grey-level non-uniformity and run-length non-uniformity, 4) structural patterns measures lacunarity, fractal dimension, and 5) multi-resolution spectral features.^18^ They conclude from this review that multiparametric texture features may be more effective in predicting breast cancer than single group features. To address use of more comprehensive summaries of these features since their review in 2016, we conducted a systematic review of published studies.

We aim to summarize the methods used to classify mammographic breast parenchymal features in relation to prediction of future breast cancer, the time from mammogram to diagnosis of breast cancer, and the analysis of data from either one or both breasts (averaged or assessed individually). We then identify gaps in evidence to prioritize future studies and speed us to better support precision prevention of breast cancer.

## Methods

### Eligibility Criteria

#### Population

We considered all studies of adult women (at least eighteen years old) involving original data. Abstract-only papers, review articles, and conference papers were excluded. Intervention: We included studies measuring at least one non-density mammographic feature. A study had to explicitly define the mammographic features used to be included. Studies that did not do this, such as those which used a deep model, were excluded.

#### Outcomes

Our primary outcome of interest was risk of breast cancer, including both invasive and in situ cancers. Risk of breast cancer was required to be dichotomized (yes/no), and analysis of other risks (e.g., risk of interval vs. screen-detected cancer) were not included.

Studies examining the association between mammographic features and other risk factors were excluded to narrow the scope of our paper.

Only studies available in English were included. Additionally, only studies published from 2016 onwards were included to avoid overlap with previous reviews.

### Information Sources

The published literature was searched using strategies designed by a medical librarian for the concepts of breast density, mammography, and related synonyms. These strategies were created using a combination of controlled vocabulary terms and keywords, and were executed in Medline (Ovid) 1946-, Embase.com 1947-, CINAHL Plus 1937-, Scopus 1823-, Cochrane Library (including CENTRAL), and Clinicaltrials.gov. Results were limited to English using database-supplied filters. Letters, comments, notes, and editorials were also excluded from the results using publication type filters and limits.

### Search Strategy

An example search is provided below (for Embase).

(‘breast density’/exp OR ((breast NEAR/3 densit*):ti,ab,kw OR (mammary NEAR/3 densit*):ti,ab,kw OR (mammographic NEAR/3 densit*):ti,ab,kw)) AND (‘mammography’/deOR mammograph*:ti,ab,kwOR mammogram*:ti,ab,kwOR mastrography:ti,ab,kwOR ‘digital breast tomosynthesis’:ti,ab,kwOR ‘x-ray breast tomosynthesis’:ti,ab,kw)NOT(‘editorial’/it OR ‘letter’/it OR ‘note’/it) AND [english]/lim

The search was completed for the first time on September 9, 2020, and was run again on October 14, 2021 to retrieve citations that were published since the original search. The second search was dated limited to 2020-present (October 2021). Full search strategies are provided in the appendix.

### Selection Process

Two reviewers (AA, CS) worked independently to review the titles and abstracts of the records. Next, the two reviewers independently screened the full text of the articles that they did not reject to determine which were eligible for inclusion. Any disagreements of which articles to include were resolved by consensus. Two reviewers (AA, YC) then went through this subset independently and excluded the ones without explicitly defined features.

### Data Collection Process

We created a data extraction sheet which two reviewers (AA, YC) used to independently extract data from the included studies. Disagreements were resolved by a third reviewer. If included studies were missing any desired information, any additional papers from the works cited, such as previous reports, methods papers, or protocols, were reviewed for this information.

### Data Items

Any estimate of risk of breast cancer was eligible to be included. Risk models could combine multiple texture features or examine texture features individually. Predictive ability could be evaluated using an area under the curve, odds ratio, matched concordance index, hazard ratio, or p value. No restrictions on follow-up time were placed. For studies that reported multiple risk estimates, we prioritized the area under the curve with the most non-mammogram covariates included from the validation study if applicable. If the study did not report an area under the curve, we listed the primary models which were discussed in the results section of the paper.

We collected data on:

the report: author, publication year

the study: location/institution, number of cases, number of controls

the research design and features: lapsed time from mammogram to diagnosis

the mammogram: machine type, mammogram view(s), breast(s) used for analysis

the model: how density was measured, number of texture features extracted, types of texture features extracted, whether feature extraction was machine or human, whether all features were used in analysis, how features for analysis were chosen, non-mammogram covariates included, prediction horizon

### Risk of Bias

The objective of this review is to summarize the methods and analysis techniques used to assess parenchymal texture features and risk of breast cancer rather than to quantitatively synthesize the results of the studies. Therefore, a risk of bias assessment, while typically performed in a systematic review, would not serve the objective and was not performed.

### Human subjects

This study did not involve human subjects and therefore oversight from an Institutional Review Board was not required.

### Registration and protocol

This review was not registered and a protocol was not prepared.

## Results

The search and study selection process is shown in Figure 1. A total of 11,111 results were retrieved from the initial database literature search and imported into Endnote. 11 citations from ClinicalTrials.gov were retrieved and added to an Excel file library. After removing duplicates, 4,863 unique citations remained for analysis. The search was run again in October 2021 to retrieve citations that were published since the original search. A total of 1,633 results were retrieved and imported to Endnote. After removing duplicates, including duplicates from the original search, 466 unique citations were added to the pool of results for screening. Between the two searches, a total of 11,577 results were retrieved, and there were 5,329 unique citations.

**Figure 1:**
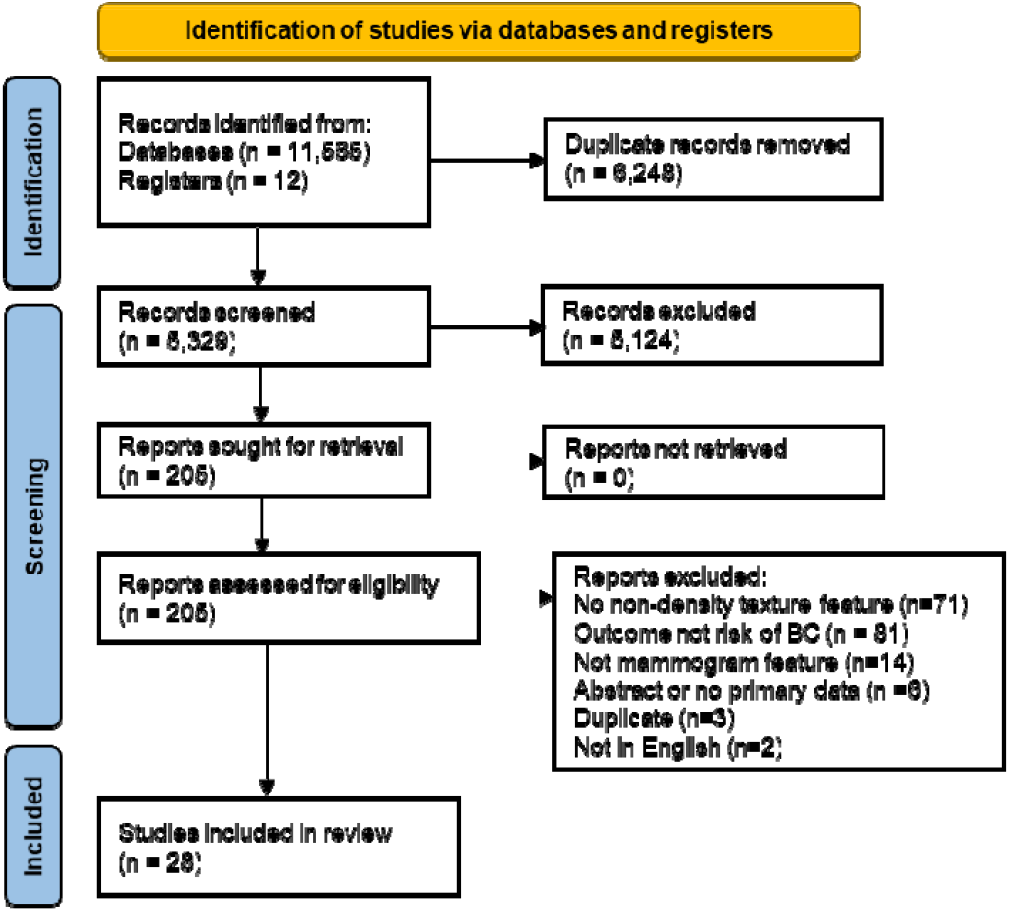
PRISMA 2020 Flow Diagram

Of the 5,329 unique citations, 5,124 were excluded based on review of title and abstract. 205 full-text reports were retrieved and assessed for eligibility by two readers. Of these, 177 were excluded for reasons such as not measuring a non-density feature, not having risk of breast cancer as an outcome, being an abstract or a duplicate paper, or not being published in English.

We identified 28 studies published since 2016 that met eligibility criteria as set out in the selection flow chart.^19-46^.

Of the 28 studies, only 7 were based on digitized analogue film images, 3 did not provide details, and the others used full field digital mammograms from Hologic, GE, or other manufacturers, or did not report details (see Table 1). The number of cases included in studies ranged from 20 up to 1900. Of the 28 studies, 8 included fewer than 100 cases.

**Table 1.**
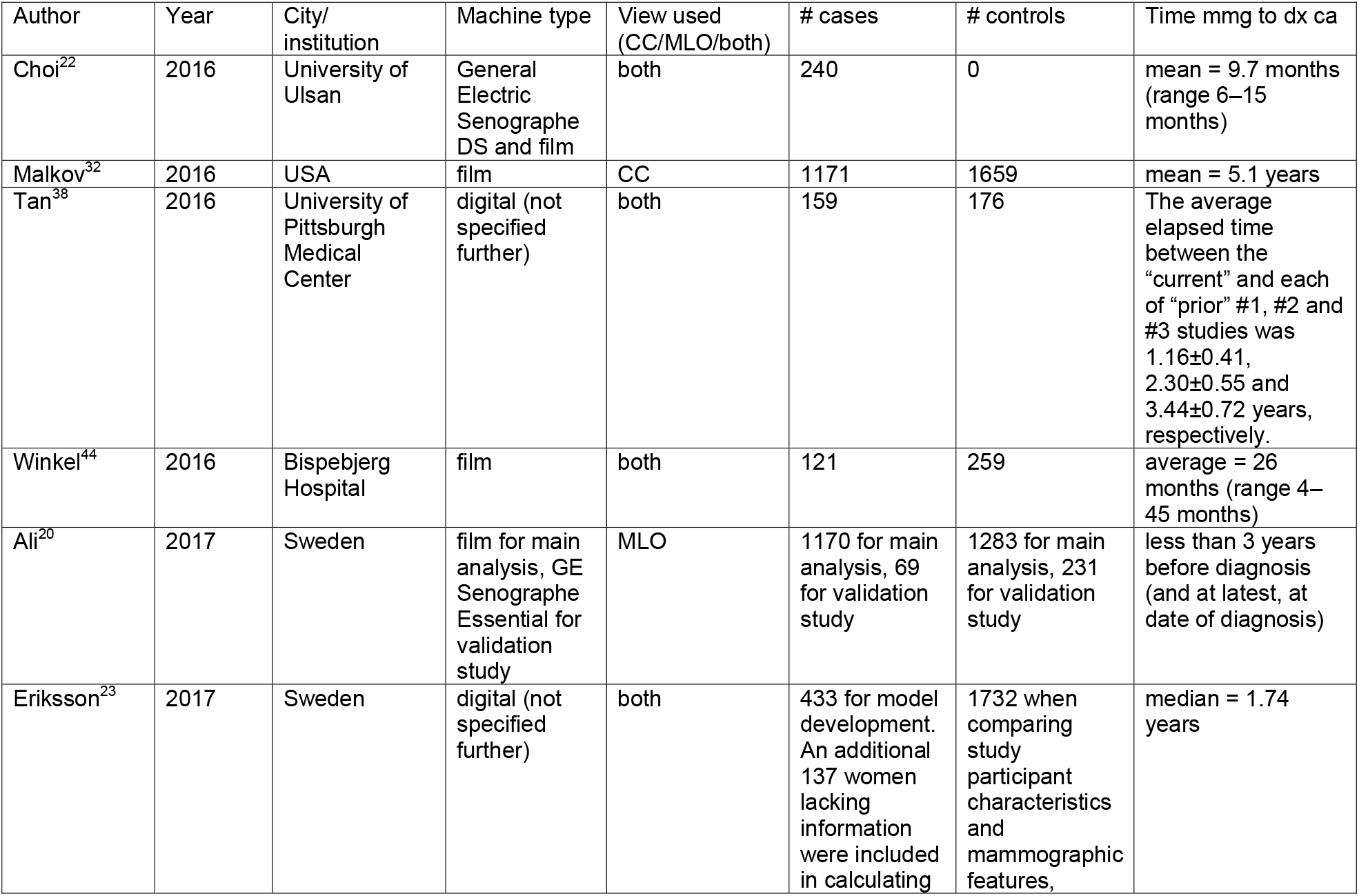
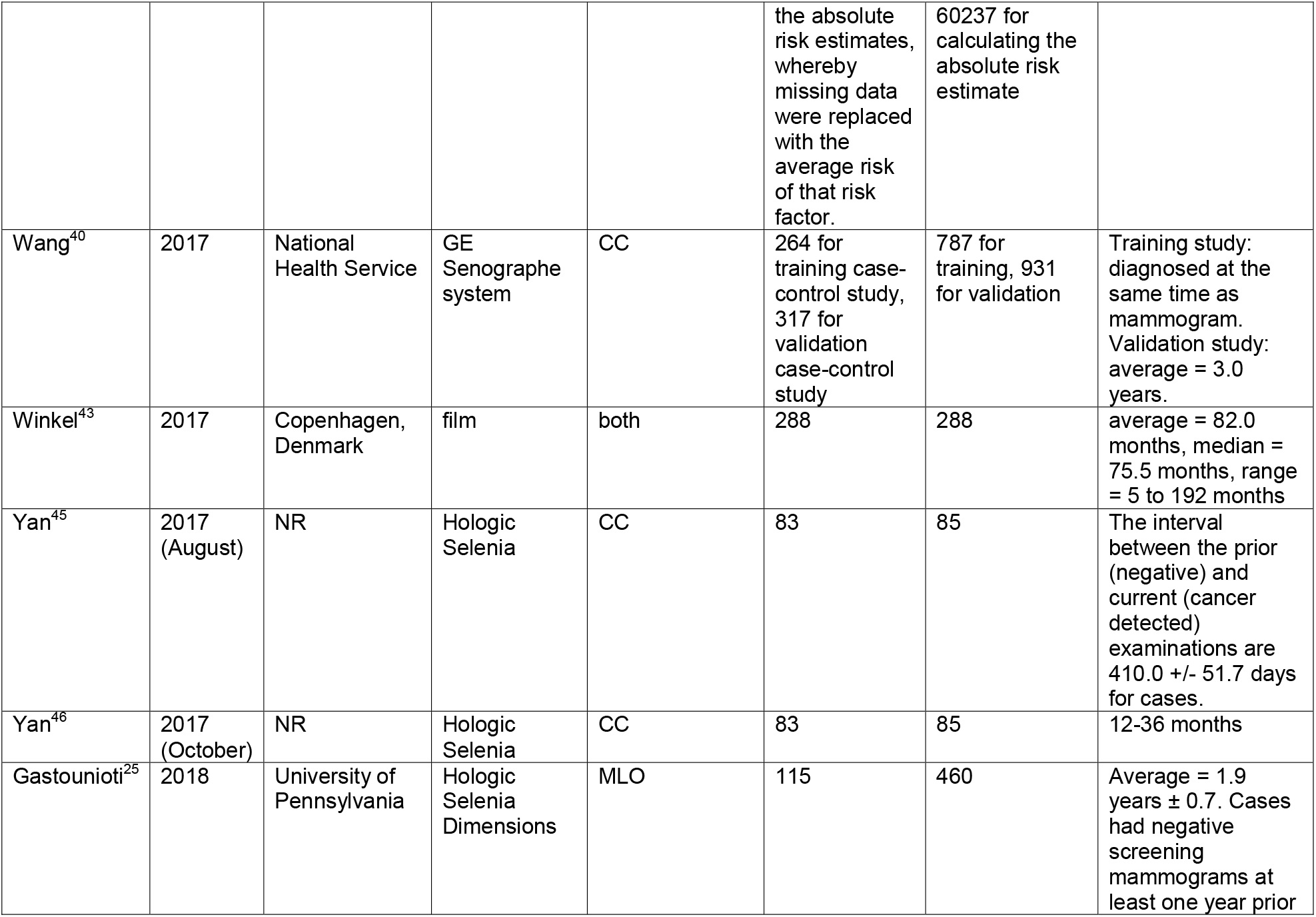
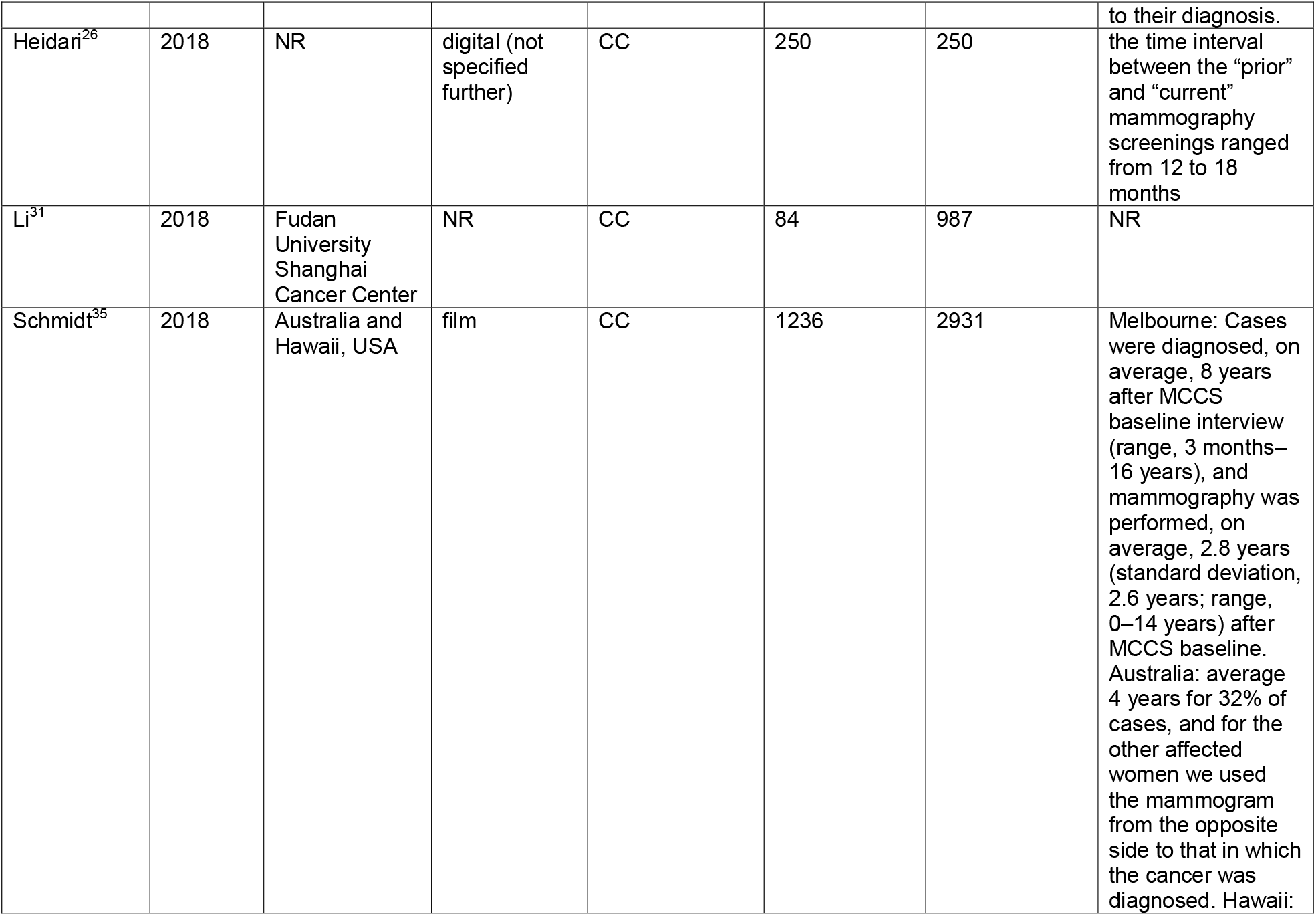
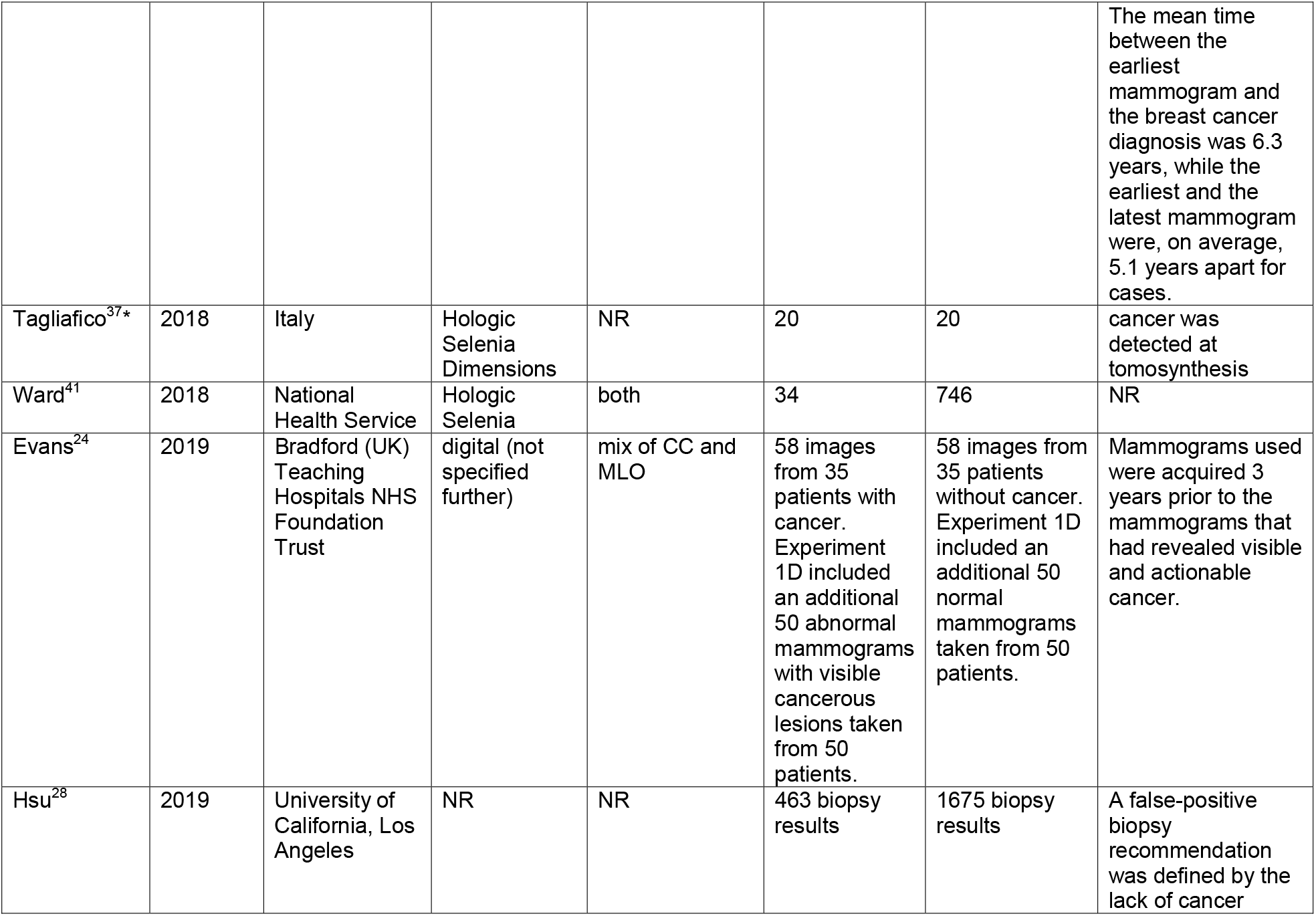
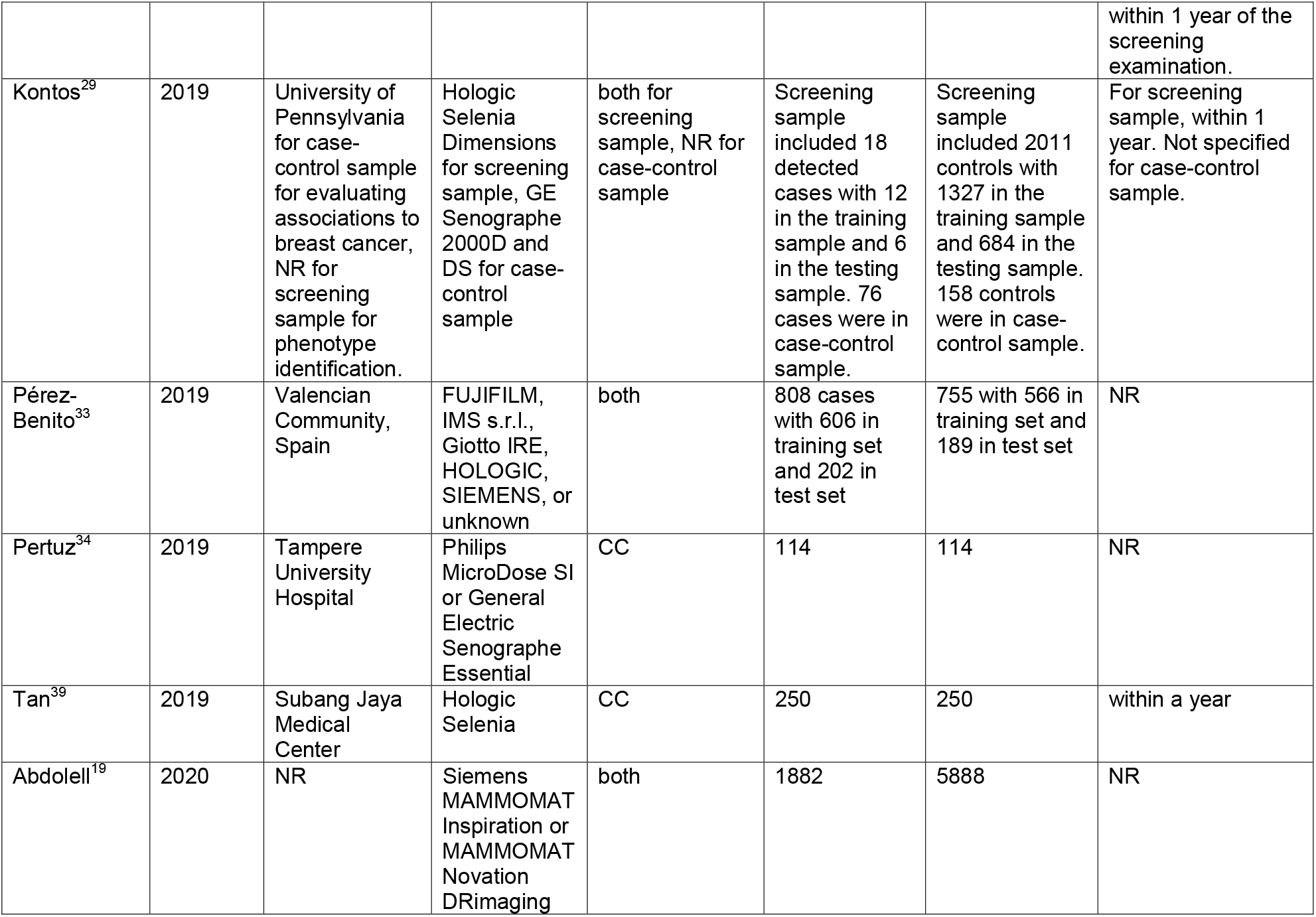
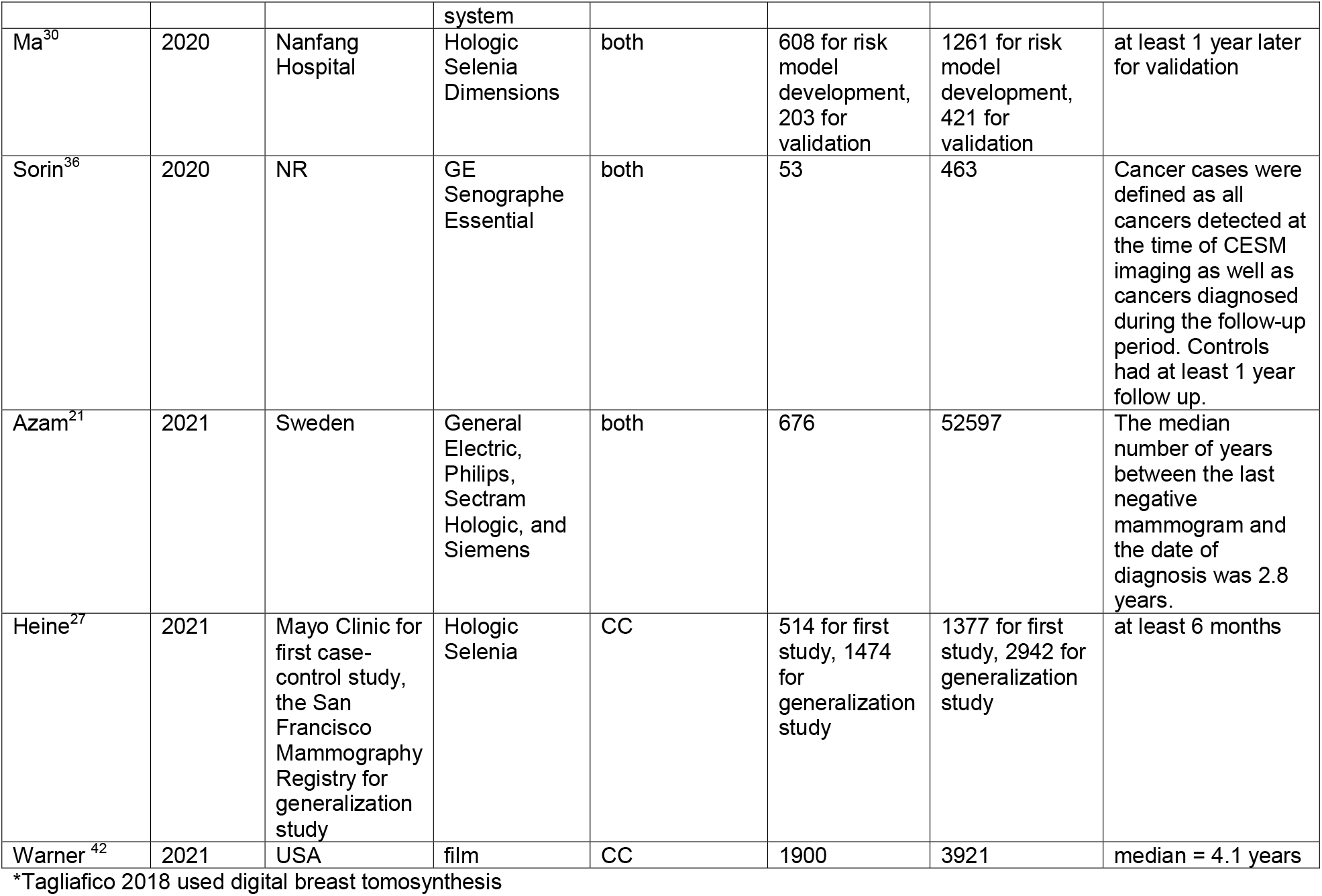
Studies of breast texture features classified from mammograms included in systematic review (sorted by year published)

The approach to assessment of breast parenchymal texture and side of body (ipsilateral or contralateral breast) varied across studies. Almost half of the studies (13 out of 28) used BIRADs as the baseline measure of density. Others used Volpara and machine-derived density or Cumulus-like approaches. Time from mammogram to diagnosis of breast cancer ranged from under 24 months on average up to 82 months.

Table 2 summarizes how the texture features were identified in the mammographic images. These methods included defined masses or calcifications^23^, or a predefined list of texture features such as 34^25^ or 44^26^ or even 112^40^ or 944^39^ initial features. Machine-driven identification of features was also reported. After machine identification, the features were reduced for analysis often based on statistical rules.

**Table 2.** Features of mammographic breast images used to assess breast texture features in addition to mammographic breast density and breast cancer risk (sorted by year).

| Author | Year | Side used (L/R/both/avg) | Density (BIRAD categories/continuous) | Number of texture features extracted (other than density) | Types of Texture features extracted (list all) | Machine or human extraction? | All texture feature used in the model (yes/no) | How features for analysis are chosen |
| --- | --- | --- | --- | --- | --- | --- | --- | --- |
| Choi | 2016 | NR | BIRADS | 8 | normal appearing tissue, benign-appearing calcification, mass, calcification, architecture distortion, focal asymmetry | human | yes | N/A |
| Malkov | 2016 | avg | Cumulus and custom software comparable to Cumulus | 46 | first- and second-order features, Fourier transform, and fractal dimension analysis | machine | no | AUCs for each feature individually given |
| Tan | 2016 | both | computer-aided detection scheme | 158 | mammographic density, structural similarity, and texture based image features | machine | no | stepwise regression analysis |
| Winkel | 2016 | both | BIRADS | 1 | Tabár classification of parenchymal patterns | human | yes | N/A |
| Ali | 2017 | contralateral for cases, random side chosen for controls | Cumulus and automated measure of area PD | 13 | spatial organisation of dense vs. fatty regions of the breast | machine | yes for AUC given, no for further analysis | stepwise selection procedure |
| Eriksson | 2017 | both | BIRADS and STRATUS | 2 | calcifications and masses | machine | yes | N/A |
| Wang | 2017 | Training: contralateral for cases and the left for controls. Validation: contralateral for cases and the same side for controls. | Volpara | 112 | features based on a grey-level co-occurrence matrix, neighborhood grey-tone difference matrix, form and shape of breast boundary, run-length, and grey-level size zone matrix, and statistical moments of pixel values | machine | no | selected from training set using least absolute shrinkage and selection operator |
| Winkel | 2017 | both | BIRADS and Cumulus-like approach | 1 | Tabár classification of parenchymal patterns | human | yes | N/A |
| Yan | 2017 (August) | both | BIRADS | 148 | bilateral mammographic tissue asymmetry maximum features | machine | no | WEKA data mining and machine learning software package |
| Yan | 2017 (October) | both | mutual threshold | 220 | asymmetry, mean and maximum features | machine | no | WEKA data mining and machine learning software package |
| Gastounioti | 2018 | avg | BIRADS, LIBRA, and Quantus | 34 | anatomically-oriented texture features | machine | no | Identified pairs of features with absolute Pearson correlation greater than 0.90 and for each pair removed the feature with the lowest variability in terms of its interquartile range. Starting from the remaining features, elastic net regression with nested cross-validation was used to build a parsimonious logistic regression model with the most discriminatory subset of covariates. |
| Heidari | 2018 | both | BIRADS | 44 | bilateral asymmetry of mammographic tissue density distribution | machine | no | locally preserving projection based feature combination algorithm |
| Li | 2018 | both | AutoDensity | 1 | breast area | machine | yes | N/A |
| Schmidt | 2018 | avg | Cumulus | 20 | gray-level co-occurrence matrix textural features | machine | yes | N/A |
| Tagliafico | 2018 | avg | BIRADS | 104 | radiomics features including skewness, energy, entropy, kurtosis, 90percentile and dissimilarity | machine | no | selected to reduce the risk of over-fitting and according to features previously used to associate breast parenchymal patterns with cancer risk |
| Ward | 2018 | both | Volpara | 1 | patterns of parenchymal tissue | human | yes | N/A |
| Evans | 2019 | both | scale similar to BIRADS used | 1 | non-localizable global gist signal | human | yes | N/A |
| Hsu | 2019 | NR | BIRADS | 5 | presence of lump, mass, calcification, architecture distortion, asymmetry | human | no | presence of lump included in final model. mass, calcifications, architecture distortion, asymmetry examined individually with PPV values given. |
| Kontos | 2019 | avg | BIRADS and LIBRA | 29 | phenotypes of mammographic parenchymal complexity based on four main types of features: histogram, co-occurrence, run-length, and structural | machine | no | excluded features with extremely low variation and those with extreme skewness |
| Pérez-Benito | 2019 | contralateral | DMScan | 23 | geometrical features and a global feature based on local histograms of oriented gradients | machine | yes | N/A |
| Pertuz | 2019 | contralateral for cases, right for controls | Cumulus-like approach | 37 | parenchymal features including computational features and imaging parameters related to the mammographic system (compressed breast thickness, compression force, X-ray tube voltage peak and target-filter combination) | machine | yes | N/A |
| Tan | 2019 | contralateral | Volpara | 944 | grey-level co-occurrence matrix features, structural/pattern measures, grey-level intensity/histogram features, run-length features, and multiresolution/spectral features | machine | no | stepwise regression analysis |
| Abdolell | 2020 | NR | Densitas | 1 | breast volume | machine | yes | N/A |
| Ma | 2020 | both | BIRADS | 1 | normalized average glandular dose | machine | yes | N/A |
| Sorin | 2020 | both | BIRADS | 1 | background parenchymal enhancement | human | yes | N/A |
| Azam | 2021 | both | STRATUS | 1 | microcalcification clusters | machine | yes | N/A |
| Heine | 2021 | avg | Volpara | 4 | variation measures | machine | no | the two variants of V produced similar findings so only one was discussed in the results |
| Warner | 2021 | both | Cumulus and automated<br>computer<br>algorithm | 1 | gray-scale variation | machine | yes | N/A |

We next assessed the value added from addition of texture features to prediction models for breast cancer. Many papers only reported on the association of texture with risk of breast cancer using an odds ratio or relative risk. These were often contemporaneous with diagnosis (measured on contralateral breast).^33,40,47-49^ Model building details and results were not routinely reported. Table 3 gives the AUC for a baseline model without texture and then the value for the model with texture when these were reported separately. Studies were not comparable across design, time horizon, and baseline models. Hence, we did not proceed to a numerical summary such as meta-analysis of AUC values. However, within studies we saw that those that reported concordance for models primarily reported performance of models with MD and then MD plus texture features. In these studies, we saw an increase in reported concordance when texture features were added. Addition of parenchymal features often increased the AUC by 0.05 though Pertuz noted an increase in AUC from 0.609 to 0.786 when using texture features in the contralateral breast at the time of cancer diagnosis.^48^ Furthermore as noted in Table 2, there was substantial variation in the number of texture features included and the method of their identification for inclusion (human defined or machine identified).

**Table 3.** Analytical models used for breast cancer risk with addition of breast texture features in addition to mammographic breast density (sorted by year).

| Author | Year | non mammogram covariates included (e.g., age, parity, etc) | Prediction horizon (<5?/5/10yr) | AUC (baseline model) | Overall AUC (with texture features added) |
| --- | --- | --- | --- | --- | --- |
| Malkov | 2016 | adjusted for age, body mass index, first-degree family history, percent density, study | NR | 0.617 | 0.621 |
| Tan | 2016 | none | NR | NR | 0.730 |
| Winkel | 2016 | adjusted for age | NR | BI-RADS density = 0.63 | 0.65 |
| Ali | 2017 | age, BMI, density, hormone replacement therapy status, parity, age at first birth | NR | 0.687 for apparent, 0.634 for honest | 0.703 for apparent, 0.643 for honest |
| Eriksson | 2017 | percentage mammographic density, age at mammography, BMI, family history of breast cancer, HRT use | 2 years (for main model) and 3 years (relative risks given) | 0.64 | 0.71 with density and interaction between percentage density and masses also included |
| Wang | 2017 | adjusted for age, BMI | NR | mC = 0.57 | mC = 0.58 |
| Winkel | 2017 | adjusted for birth year, age at false-positive-screen, invitation round at false-positive-screen | NR | BI-RADS density = 0.65, percentage mammographic density = 0.62 | 0.63 |
| Yan | 2017 (August) | none | next sequential mammography screening | NR | 0.816 |
| Yan | 2017 (October) | none | next sequential sequencing | NR | 0.830 |
| Gastounioti | 2018 | density, BMI, age | NR | 0.62 | 0.67 |
| Heidari | 2018 | none | 12 to 18 months | NR | 0.68 |
| Schmidt | 2018 | adjusted for age, BMI | NR | percent mammographic density = 0.620 | 0.662 |
| Tagliafico | 2018 | none | NR | NR | 0.567 |
| Evans | 2019 | none | 3 years | NR | 0.54 |
| Hsu | 2019 | BI-RADS, density, age, race, BMI, age at first live birth, noticeable changes in breast, number of risk factors, 5-year Gail risk $\geq 1.67\%$ | NR | cv-AUC = 0.83 with BI-RADS and density only | cv-AUC = 0.84 |
| Kontos | 2019 | density, BMI | NR for AUC model. 5-year risk from Gail and Breast Cancer Surveillance Consortium models compared by phenotype. | 0.80 | 0.84 |
| Pérez-Benito | 2019 | percent density | NR | 0.560 | 0.614 |
| Pertuz | 2019 | none and with age, percent density | NR | density only = 0.609 | 0.786 |
| Tan | 2019 | none, with age only, and with age and BMI | NR | density = 0.52 | 0.68 |
| Abdolell | 2020 | age, percent mammographic density | tailored estimates of current breast cancer risk | 0.584 | 0.597 |
| Ma | 2020 | age, age at menarche, menopausal status, age at first birth, parity, family history of breast cancer, breast density | NR | 0.61 for training set and 0.56 for test set | 0.77 for training set and 0.75 for test set |
| Heine | 2021 | adjusted for study, age, body mass index and also with dense volume included | NR | volumetric breast density = 0.61 for first study and 0.59 for generalization study | $V = 0.61$, $P_1 = 0.61$, $p_1 = 0.60$ for first study. $V = 0.59$, $P_1 = 0.57$, $p_1 = 0.58$ for generalization study. |

The prediction horizon was only defined for 3 of the studies (see Table 3) and was usually the time to the next routine screening mammogram, but less than 3 years on average. Examples of several studies are summarized to give more context to the details in the tables. Eriksson ^50^ evaluated data from the KARMA cohort that followed 70,877 women for up to 3 years after baseline mammograms. Median time from the screening mammogram to breast cancer diagnosis was 1.74 years and 433 breast cancers were diagnosed. In their analysis, they used both 2- and 3-year horizons. The AUC improved from 0.64 for the model that included age, MD, and BMI to 0.71 after adding calcifications. Heidari 2018 ^26^ chose 4 features associated with asymmetry of mammogram images from a pool of 44 machine-identified features. The time horizon was 12 to 18 months, defined as time to the next screening mammogram. A machine learning classifier was built to predict breast cancer on the subsequent mammogram, reducing the features to a vector with 4 features. The AUC improved from 0.62 to 0.68.

Not all studies reported concordance statistics for their analysis. We summarized 6 studies in Table 4 that reported association measures for texture features with breast cancer.

**Table 4.** Studies without AUC (sorted by year).

| Author | Year | non mammogram covariates included (e.g., age, parity, etc) | Prediction horizon (<5?/5/10yr) | Risk other than AUC |
| --- | --- | --- | --- | --- |
| Choi | 2016 | N/A | N/A | In the minimal sign group, the most common finding was normal appearing tissue (61/78), followed by benign-appearing calcification (17/78). |
| Li | 2018 | none | NR | OR = 1.018 |
| Ward | 2018 | none | NR | There was a significant correlation between a diagnosis of cancer and nodular parenchymal pattern compared to not nodular parenchymal pattern ($p = 0.043$). |
| Sorin | 2020 | adjusted for age, family history, breast density | NR | The odds for breast cancer significantly increased with increased BPE (OR = 2.24). |
| Azam | 2021 | adjusted for BMI, baseline mammographic density, smoking status, alcohol consumption, age at menarche, age at first birth, number of children, breastfeeding duration, oral contraceptive use, menopausal hormone therapy use, family history of breast cancer | NR | Each additional microcalcification cluster was associated with 20% increased risk of breast cancer (HR = 1.20). Women with $\geq 3$ microcalcification clusters had an overall 2-fold increased risk of breast cancer compared to women with no clusters (HR = 2.17). |
| Warner | 2021 | adjusted for age, fasting status, time of blood draw, body mass index, menopausal status, hormone therapy use, mammography read batch and also with either percent mammographic density or automated percent density | NR | V was positively associated with breast cancer risk (OR = 1.15 with percent mammographic density, 1.18 with automated percent density). |

## Discussion

We identified 28 studies that evaluated the value of adding texture features to predict future breast cancer incidence or reported the association between these features and risk. Of these, 7 were based on film images, 3 did not report details, and the others used digital mammograms. Among these, we observed 5 studies using measures of texture that were measured or defined at time of cancer diagnosis and from the contralateral breast. However, 5 studies did not report a time horizon for the interval from mammogram to cancer diagnosis. Those studies evaluating future risk are limited to less than 3 years, on average. When evaluated as a contributor to risk of future breast cancer diagnosis, the common measure of discrimination, the AUC, increases when breast texture measures are added to mammographic breast density. We did not identify any study using mammography texture features to predict 5- or 10-year risk of breast cancer that would be sufficient to guide prevention.^6,51-54^

There is substantial variation in the methods used for defining and summarizing the measures of texture. No consistent approach is used to reduce the large number of predefined features, or machine-identified features, to a subset or summary for analysis. Given this variation, we did not combine data across studies but note that comparison within studies shows that texture features are related to breast cancer incidence and improved concordance or AUC. Texture features are important contributors to breast cancer risk beyond mammographic breast density.

In the 2016 review, Gastounioti et al. reviewed features of automated parenchymal texture analysis in relation to breast cancer risk.^18^ This included details of methods to classify texture in mammographic images and its contribution to discrimination in case-control studies. Based on the review, they concluded that further research including large prospective studies is needed to establish the predictive value of parenchymal texture for ultimate inclusion in breast cancer risk prediction models. Such prediction models that extend to 5 and 10 years then require external validation to support clinical risk management.

Regarding our methods for this updated systematic review, we do not use risk of bias given the inconsistent approaches and the lack of quantitative data to estimate a summary measure of change in AUC. There are several limitations with the current review. Heterogeneity of the data did not allow for a meta-analysis. Additionally, systematic reviews are always subject to possible publication bias if all relevant studies have not been published. We used several strategies to reduce the risk of this including using a thorough search strategy designed by a medical librarian with expertise in searching for systematic reviews, and searching clinicaltrials.gov for any ongoing studies.

For clinical use to guide precision prevention we must identify both high-risk women for a range of risk reduction strategies^6^ and low-risk women to consider frequency of screening.^55^ From this systematic literature review we identify gaps in evidence to prioritize future studies. They include: i) details on prediction horizon for risk of breast cancer; ii) other statistical approaches might also be used to assess risk of breast cancer from time of image acquisition that include the texture features to the diagnosis of breast cancer such as survival analysis.

## Conclusion

Despite current limitations in the literature, the more widespread use of digital mammography and availability of digital images including parenchymal features offer growing opportunity to more uniformly assess image texture features and incorporate these into risk prediction models that can improve risk classification and risk prediction for women.

## Data Availability

All data produced in the present study are available upon reasonable request to the authors

## Appendix Complete Search Strategies

### Search strategies designed and executed by Angela Hardi, MLIS

#### Embase.com

=3,919 results on 9/9/2020 (Limited to English; editorials, letters, and notes excluded from results)

**Updated search** (date limited to 2020-present): 602 on 10/14/2021

(‘breast density’/exp OR ((breast NEAR/3 densit*):ti,ab,kw OR (mammary NEAR/3 densit*):ti,ab,kw OR (mammographic NEAR/3 densit*):ti,ab,kw)) AND (‘mammography’/de OR mammograph*:ti,ab,kw OR mammogram*:ti,ab,kw OR mastrography:ti,ab,kw OR ‘digital breast tomosynthesis’:ti,ab,kw OR ‘x-ray breast tomosynthesis’:ti,ab,kw) NOT (‘editorial’/it OR ‘letter’/it OR ‘note’/it) AND [english]/lim

#### Ovid Medline All

= 2694 results on 9/9/2020 (Limited to English; editorials, comments, and letters excluded)

**Updated search** (date limited to 2020-present): 440 results on 10/14/2021

(Breast Density/ OR (breast adj3 densit*).ti,ab. OR (mammary adj3 densit*).ti,ab. OR (mammographic adj3 densit*).ti,ab.) AND (Mammography/ OR mammograph*.ti,ab. OR mammogram*.ti,ab. OR mastrography.ti,ab. OR “digital breast tomosynthesis”.ti,ab. OR “x-ray breast tomosynthesis”.ti,ab.) NOT (comment.pt. OR editorial.pt. OR letter.pt.)

#### CINAHL Plus

=978 results on 9/9/2020; (Limited to English and these publication types: Clinical Trial, Corrected Article, Journal Article, Meta Analysis, Meta Synthesis, Practice Guidelines, Proceedings, Protocol, Randomized Controlled Trial, Research, Review, Systematic Review)

**Updated search** (dated limited to 2020-present): 135 results on 10/14/2021

((MH “Breast Tissue Density”) OR AB(breast N3 densit*) OR TI(breast N3 densit*) OR AB(mammary N3 densit*) TI(mammary N3 densit*) OR AB(mammographic N3 densit*) OR TI(mammographic N3 densit*)) AND ((MH “Mammography”) OR AB(mammograph*) OR TI(mammograph*) OR AB(mammogram*) OR TI(mammogram*) OR AB(mastrography) OR TI(mastrography) OR AB(“digital breast tomosynthesis”) OR TI(“digital breast tomosynthesis”) OR AB(“x-ray breast tomosynthesis”) OR TI(“x-ray breast tomosynthesis”))

#### Scopus

=3,162 results on 9/9/2020 (Limited to English; editorials, notes, letters, and book chapters excluded from results)

**Updated search** (date limited to 2020-present): 423 results on 10/14/2021

TITLE-ABS ((breast W/3 densit*) OR (mammary W/3 densit*) OR (mammographic W/3 densit*)) AND TITLE-ABS (mammograph* OR mammogram* OR mastrography OR “digital breast tomosynthesis” OR “x-ray breast tomosynthesis”) AND (EXCLUDE (DOCTYPE, “no”) OR EXCLUDE (DOCTYPE, “ch”) OR EXCLUDE (DOCTYPE, “le”) OR EXCLUDE (DOCTYPE, “ed”)) AND (LIMIT-TO (LANGUAGE, “English”))

#### Cochrane Library

=358 results on 9/9/2020 (1 Cochrane Protocol and 357 results from CENTRAL Trials)

**Updated Search** (date limited 2020-present in CENTRAL Trials) = 32 results on 10/14/2021

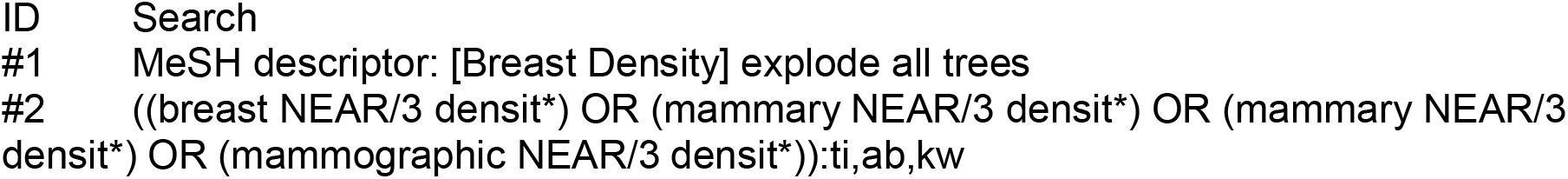
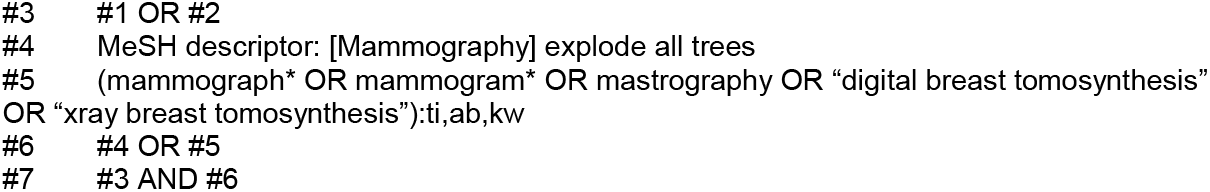

#### ClinicalTrials.gov

= 11 results (searched the “Other terms” field) on 9/9/2020

**Updated Search** = 12 results on 10/14/2021 (1 new result, added to the Excel library) (“breast density” OR “mammary density”) AND (mammograph* OR mammogram* OR mastrography OR “digital breast tomosynthesis” OR “x-ray breast tomosynthesis”)

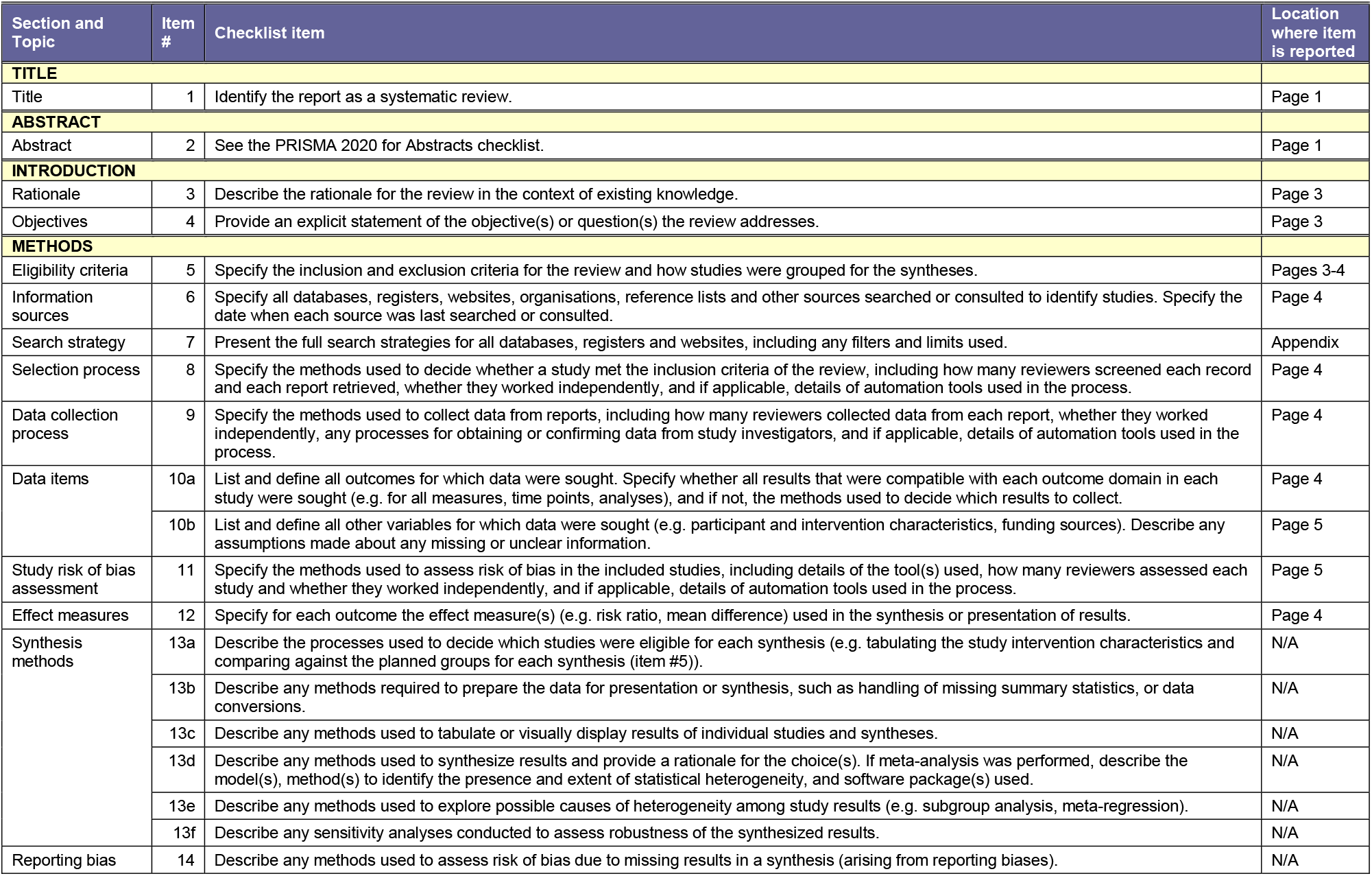
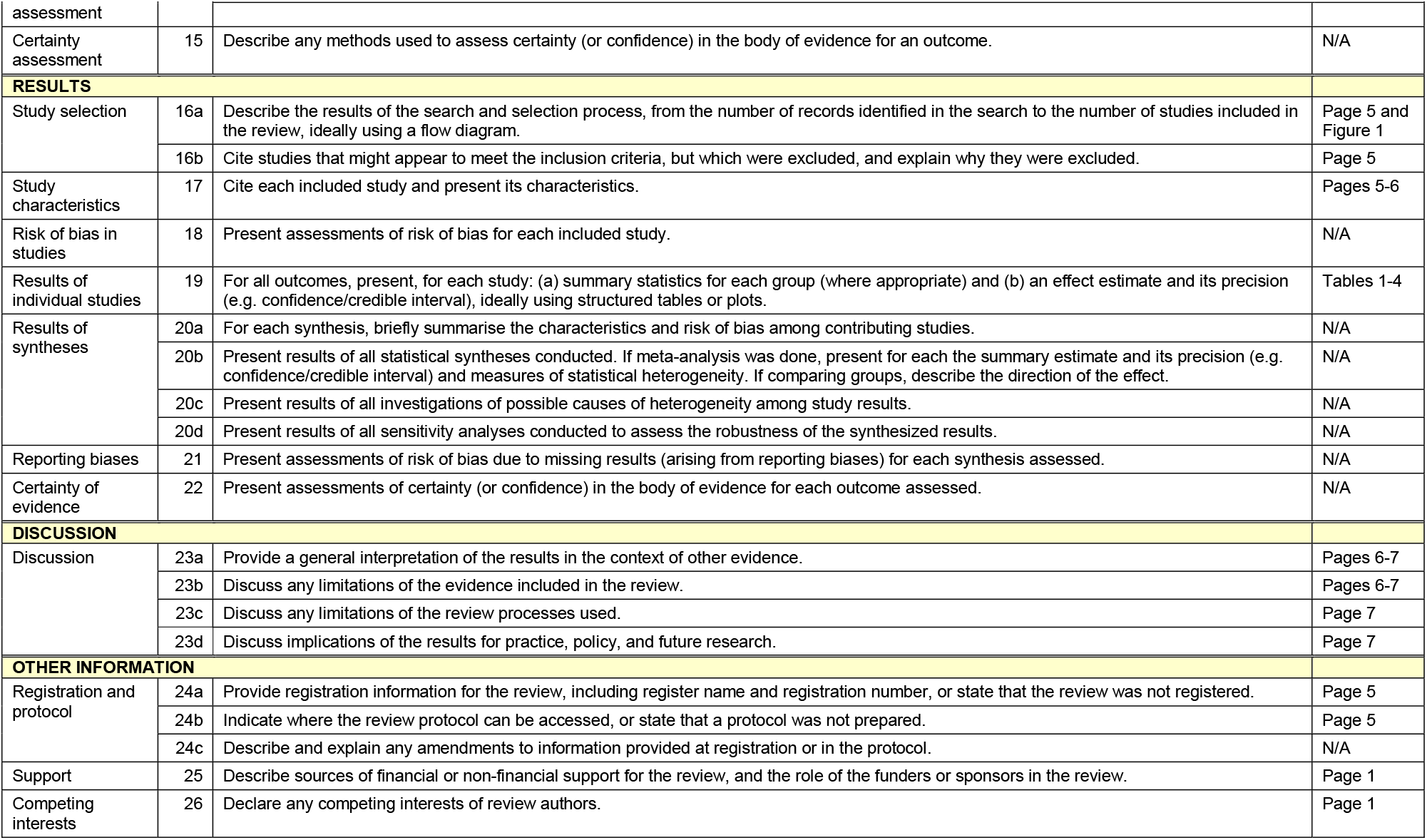
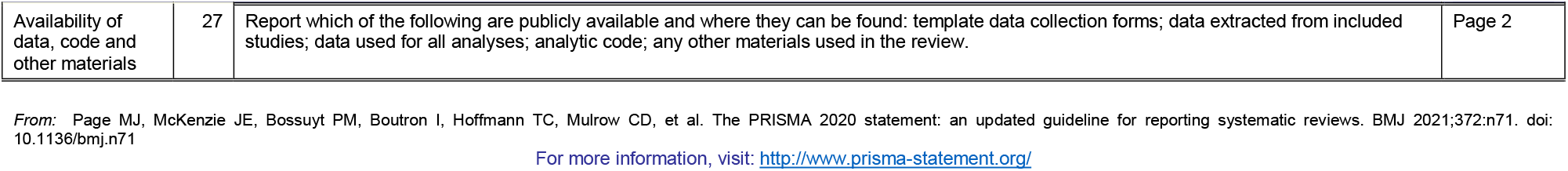

## Notes

**Funding statement** This work was supported by Breast Cancer Research Foundation grant number (BCRF 21-028), and in part by NCI (R37 CA256810).

### Competing Interest Statement

The authors have declared no competing interest.

### Funding Statement

This study was funded by Breast Cancer Research Foundation grant number (BCRF 21-028), and in part by NCI (R37 CA256810).

### Summary of Updates

Author affiliation corrected.

